# Long-term effects of mass screening for latent and active tuberculosis in the Marshall Islands

**DOI:** 10.1101/2021.03.03.21252859

**Authors:** Romain Ragonnet, Bridget M Williams, Angela Largen, Joaquin Nasa, Tom Jack, Mailynn K Langinlur, Eunyoung Ko, Kalpeshsinh Rahevar, Tauhid Islam, Justin Denholm, Ben J Marais, Guy B Marks, Emma S McBryde, James M Trauer

**Author notes:** Corresponding author: Dr Romain Ragonnet, Monash School of Public Health and Preventive Medicine, 553 St Kilda Road Melbourne VIC 3004. **Authors’ contributions:** RR, BMW, TI and JMT designed the study. AL, JNJ, TJ and MKL contributed to collecting and processing the data used to inform the model. RR, BMW, ESM and JMT designed the mathematical model. RR and JMT implemented the code. RR conducted the experiments. All authors contributed to the interpretation of the findings. RR wrote the first version of the manuscript. All authors contributed to the final version of the manuscript. **Sources of support:** This work was funded by the Australian National Health and Medical Research Council (Project Grant APP1144570 and Fellowship Grant APP1142638) and the Western Pacific Regional Office of the World Health Organization.

## Abstract

**Rationale:** The Marshall Islands implemented ambitious population-based screening programs for latent and active tuberculosis in 2017 and 2018. These interventions’ long-term effects remain to be estimated.

**Objectives:** To predict the long-term impact of the previous interventions and identify strategies to drive tuberculosis towards elimination.

**Methods:** We built a transmission model of tuberculosis informed by local data to capture the epidemic’s historical dynamics. We used the model to project the future epidemic trajectory following the screening interventions, as well as considering a counterfactual scenario with no intervention. We also simulated future scenarios including periodic interventions similar to those previously implemented, to assess the feasibility of reaching the End TB Strategy targets and tuberculosis pre-elimination.

**Measurements and Main Results:** The 2017-2018 screening activities were estimated to have reduced tuberculosis incidence and mortality by more than one third in 2020, and are predicted to achieve the End TB Strategy milestone of 50% incidence reduction by 2025 compared to 2015. Interventions had a considerably greater impact when individuals were also screened for latent infection than active case finding alone. Such combined programs implemented at the national level could achieve tuberculosis pre-elimination by 2035 if repeated every two years, and around 2045 if repeated every five years.

**Conclusions:** We predict that it is possible to achieve tuberculosis pre-elimination by 2035 in the Marshall Islands through periodic repetition of the same ambitious interventions as those previously implemented. Including latent infection testing in active screening activities will be a critical pillar for achieving these ambitious goals.

## Introduction

Every year, around ten million people develop tuberculosis (TB) disease globally (1). The causative agent of TB (*Mycobacterium tuberculosis, M*.*tb*) was first identified in 1882 – and yet more than 130 years later, TB is estimated to be the world’s leading cause of death from a single infectious agent (1). The availability of effective vaccines and treatments has not been sufficient to eliminate TB. Consequently, disease elimination is not expected in the coming decades without dramatic changes to current TB control approaches.

Improving case detection is a promising way to progress the global TB response because millions of diseased individuals are undetected (1). Active case finding (ACF) activities can identify significant numbers of individuals with active disease who may not have been detected otherwise or may only have been detected after substantial delays. Recent large-scale randomised trials of ACF interventions in high burden settings have demonstrated the efficacy of these approaches and generated great hope for dramatically improved control by reducing the pool of infectious individuals (2,3).

Screening and treatment of latent TB infection (LTBI) can also be included in these ambitious programs to prevent future TB reactivation in infected individuals. Large-scale cluster-randomised trials of preventive treatment were conducted in the late 1950s in Alaska, Greenland, and Tunisia, all demonstrating significant TB incidence reductions (4–6). However, a more recent trial conducted in a South African mining population with a high prevalence of HIV was not successful at reducing TB burden (7). This demonstrates that the effectiveness of prophylaxis programs depends on the characteristics of the population in which the intervention is implemented (8), and that it is difficult to anticipate the effects of future interventions by extrapolating observations made in other settings.

Ambitious population-level screening programs were recently completed in the Republic of the Marshall Islands (RMI), an archipelago nation in the western Pacific. RMI is a high TB burden country with an estimated TB incidence approaching 500 cases-per-100,000-persons-per-year in 2019, according to the World Health Organization (WHO) (9). The screening activities were conducted in the two most populous islands of RMI – Ebeye and Majuro – together representing nearly three-quarters of the national population. In 2017, a first ACF intervention aimed to screen the entire adult population (those aged 15 years or older) of Ebeye Island for active TB disease and treat those with suspected disease. The following year, an even more comprehensive intervention was conducted on Majuro Atoll to screen individuals of all ages for both latent and active TB and provided appropriate treatment for both these conditions. These programs have enabled the identification and treatment of a large number of infected individuals. It is now critical to estimate the long-term effects of these interventions and identify effective follow-up approaches that would sustain significant TB incidence reductions in RMI.

In this modelling study, we incorporated local data collected during the interventions into a transmission dynamic model of TB to assess whether RMI could achieve the End TB Strategy targets and pre-elimination under the current strategy, as well as under various future intervention scenarios.

## Methods

### Overall approach

We used a deterministic compartmental model to simulate *M*.*tb* transmission in RMI using a similar approach to previously published studies (10–12). After calibrating the model to local data using Bayesian techniques, we projected the long-term effect of the large-scale LTBI and TB screening activities undertaken in 2017 and 2018. We then simulated periodic interventions similar to those previously implemented and covering the entire country at regular intervals. We assessed prospects for reaching the End TB Strategy milestones and targets and the pre-elimination incidence threshold (see Table 1).

**Table 1.** End TB Strategy targets and milestones and pre-elimination threshold.

| Target or milestone | Definition |
| --- | --- |
| ***TB incidence*** |  |
| 2025 milestone (21) | 50% reduction between 2015 and 2025 |
| 2035 target (21) | 90% reduction between 2015 and 2035 |
| Pre-elimination threshold<br>(15) | TB incidence below 1 case per-100,000-per-year |
| ***TB mortality*** |  |
| 2025 milestone (21) | 75% reduction between 2015 and 2025 |
| 2035 target (21) | 95% reduction between 2015 and 2035 |

### Tuberculosis model

The base model consisted of six compartments representing particular clinical statuses regarding *M*.*tb* infection or disease (Figure 1). LTBI was modelled using two sequential compartments (*E* and *L*) to capture the declining risk of disease progression over time from infection (13). Individuals progressing to active TB (*I*) were classified based on their form of disease: smear-positive pulmonary TB, smear-negative pulmonary TB, and extrapulmonary TB. The simulated population was stratified by age and location (Majuro Atoll, Ebeye Island and all other islands of RMI). More details about the model are available in the Supplementary Appendix, including a description of time-variant BCG vaccination and type 2 diabetes prevalence.

**Figure 1.**
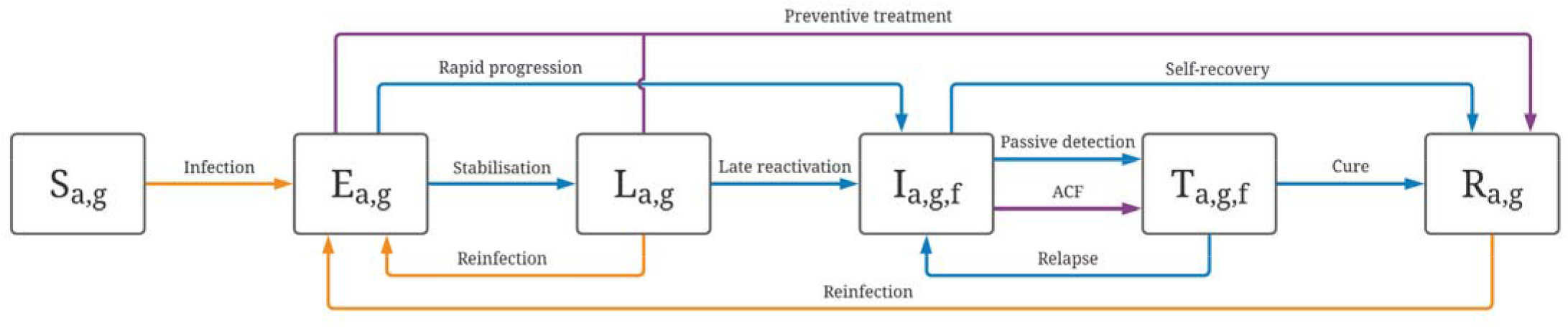
Illustration of the model structure. Boxes represent the different compartments types: susceptible (S), early latent (E), late latent (L), infectious (I), on treatment (T) and recovered (R). The subscripts indicate whether compartments are stratified by age (a), geography (g) and form of TB (f). Blue and orange arrows represent progression flows and transmission flows, respectively. The flows associated with the modelled interventions are shown in purple.

We used local programmatic data to inform the simulated detection and treatment processes. Our previously published estimates were used to inform the natural history of TB (14), and the progression rates from latent to active TB (13). We fitted model parameters using local data on population size, TB prevalence, LTBI prevalence and TB notifications while considering uncertainty around the most critical model parameters (Table 2). The code used to implement the model is publicly available on Github (https://github.com/monash-emu/AuTuMN/tree/master/apps/tuberculosis/regions/marshall_islands).

**Table 2.** Model parameters. The ranges presented for the fitted parameters correspond to the ranges used to inform the prior distributions in the adaptive Metropolis algorithm. The values of the parameters that are time-variant and/or age-specific are presented in the Supplement. *These parameters are multiplied by the fitted screening rate parameter, such that their absolute values are less significant than the relative values between the different forms of TB.

| Parameter | Value or uncertainty range | Source |
| --- | --- | --- |
| *Population characteristics* |  |  |
| Targeted total population size (2011, all RMI) | 53,158 | 2011 National Census |
| Population size at the start of simulation (1800) | 200 - 1000 | Fitted |
| Population proportions in Majuro, Ebeye and other islands (2011) | 52% / 20% / 28% | 2011 National Census |
| Proportion of contacts that occur with individuals from the same geographic group | 95% | Assumption. Remainder of contacts distributed evenly between the two other locations (2.5% each) |
| Crude birth rate | Time-variant | United Nations Population Division data for the Federated States of Micronesia |
| All-cause mortality rate | Age-specific and time-variant | United Nations Population Division |
| Type 2 Diabetes prevalence | Age-specific and time-variant | Assumed diabetes proportions in 2020 based on the age-adjusted prevalence reported by the IDF Diabetes Atlas (22):<br>0-4: 1%<br>5-14: 5%<br>15-34: 20%<br>35-49: 40%<br>50+: 70% |
| *M.tb infection and TB disease* |  |  |
| Transmission scaling factor | 0.2 - 1 | Fitted |
| Relative infectiousness (smear-positive / smear-negative / extrapulmonary TB) | 1 / 0.25 / 0 | (23,24) |
| Relative infectiousness by age | Progressive increase through childhood (Supplement) | (25,26) |
| Relative infectiousness during treatment (ref. untreated TB) | 0.08 | Based on the assumption that patients are infectious for the first two weeks of a 6-month regimen |
| Rate of stabilisation from early to late latency (age 0-4 / 5-14 / 15+) | 4.4 / 4.4 / 2 per year | (13) |
| Rate of rapid progression to active TB (age 0-4 / 5-14 / 15+) | 2.4 / 2 / 0.1 per year | (13) |
| Rate of late reactivation (age 0-4 / 5-14 / 15+) | $7e^{-9}$ / $2.3e^{-3}$ / $1.2e^{-3}$ per year | (13) |
| Uncertainty multiplier for the rates of TB progression | 0.5 - 2 | Fitted (13) |
| Relative risk of TB progression for individuals with type-2 diabetes | 2 - 5 | (27) |
| Proportion of pulmonary TB among incident TB | 85% | Adjusted to replicate observed prevalence proportions by form of TB |
| Proportion of smear-positive TB among incident pulmonary TB | 75% | Adjusted to replicate observed prevalence proportions by form of TB |
| Rate of self recovery (smear-positive TB / other forms of TB) | 0.18 - 0.29 / 0.07 - 0.21 per year | (14) |
| Rate of TB-specific mortality (smear-positive TB / other forms of TB) | 0.34 - 0.45 / 0.017 - 0.035 per year | (14) |
| Relative risk of reinfection while latently infected (ref. infection-naïve) | 0.2 - 0.5 | (28) |
| Relative risk of reinfection after recovery (ref. infection-naïve) | 0.2 - 1 | (28) |
| ***TB control*** |  |  |
| BCG vaccination coverage | Time-variant | Global Health Observatory data repository (WHO) |
| Reduced susceptibility to infection due to BCG vaccination | Age-specific | (29,30) |
| Passive TB screening rate | Varies with time and location | Fitted (see Supplement) |
| TB screening sensitivity (smear-positive / smear-negative / extrapulmonary TB) | 100% / 70% / 50% | Assumption* |
| Treatment success rate | Time-variant | WHO |
| Proportion of death among non-successful treatment | 20% | WHO |
| Average treatment duration | 6 months | Assumption |
| Active case finding rate | Varies with time, location and scenario | - |
| LTBI screening rate | Varies with time, location and scenario | - |
| Preventive treatment efficacy (intention-to-treat) | 75 - 85% | Based on completion rate during intervention |
| Relative improvement in passive screening rate following interventions | 0 - 50% | Assumption based on discussions with the national program |

### Modelled interventions

Latently infected individuals were assumed to transition to the recovered compartment on completion of LTBI treatment, at a rate defined as the product of a time-variant LTBI screening rate, the LTBI test’s sensitivity, and the efficacy of preventive treatment. The effect of ACF was modelled using a time-variant screening rate, multiplied by the sensitivity of the diagnostic test used during the ACF intervention and was assumed to move individuals from the undetected disease compartment to the treatment compartment (Figure 1). Screening rates were parameterised such that the modelled proportions of screened individuals by the end of the interventions matched those measured in the field. To capture the future effect of increased TB awareness due to the large-scale community interventions, we increased the rate of passive screening of active TB from 2018 - the endpoint of the completed intervention, considering relative improvements in the screening rate ranging from zero to 50%.

We also modelled scenarios where the community-wide interventions would be repeated periodically at the national level, every two, five or ten years, starting from 2021. We assumed a similar screening rate to that of the intervention conducted in Majuro in 2018. Finally, we estimated the effect of intensive contact tracing and preventive treatment provision in TB contacts.

## Results

Figure 2 presents the base-case model fits to local data. Posterior parameter estimates are shown in the Supplement (Table E3).

**Figure 2.**
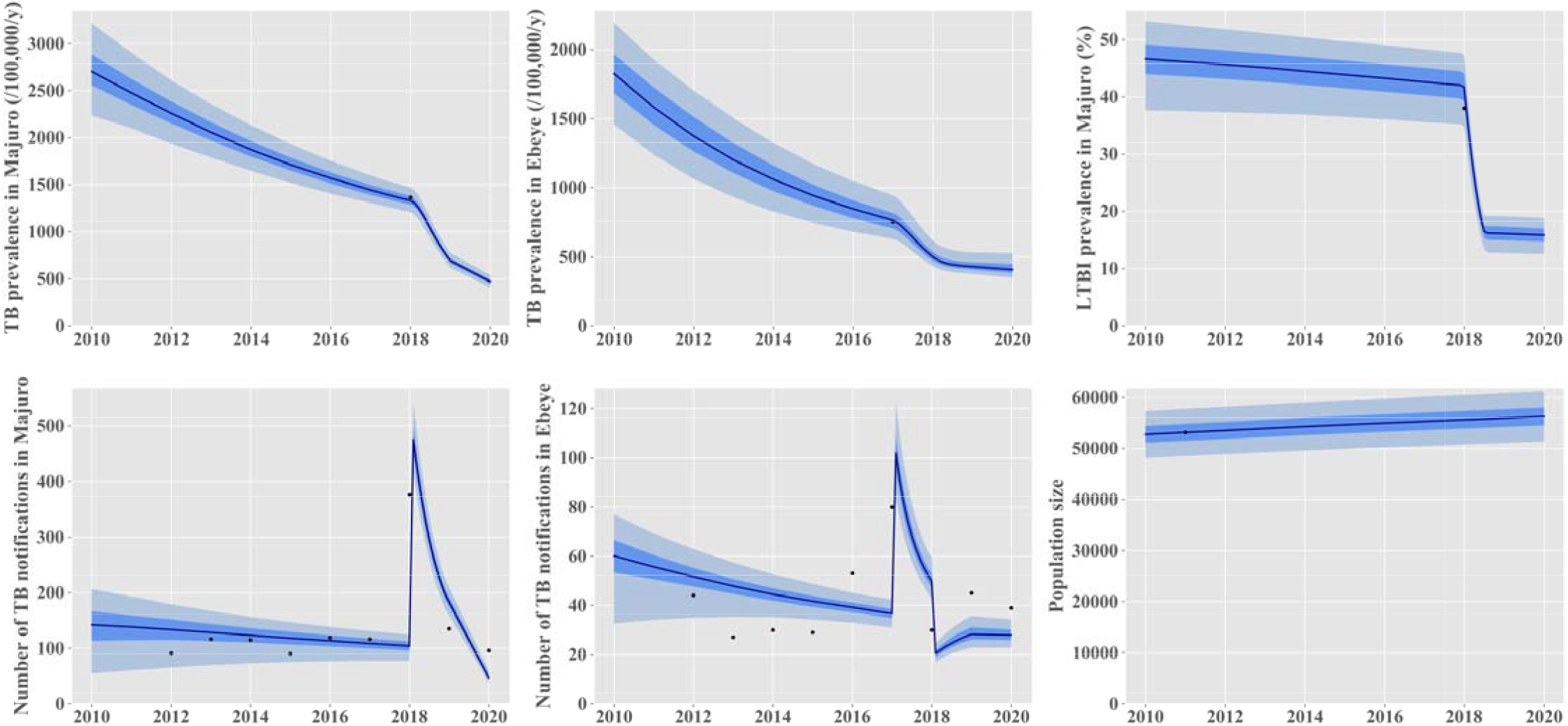
Comparison between model outputs and local data for the calibration targets. The black dots represent local empiric data. The model predictions are represented in blue as median (solid line), interquartile range (dark shade) and 95% central credible interval (light blue shade). The effect of the 2017-2018 interventions was included in these projections.

Figure 3 presents the future projected epidemic trajectories with and without the implementation of the active screening interventions. At the national level, we estimated that the interventions had reduced TB incidence by around 37% in 2020 from 447 (95% credible interval 353-548) to 281 (95% CrI 221-363) per-100,000-persons-per-year. TB mortality was predicted to have decreased by around 42% from 127 (95% CrI 108-160) to 74 (95% CrI 61-100) per-100,000-persons-per-year in 2020 due to the screening interventions. The community-wide interventions had a considerably greater effect on the epidemic in Majuro than in Ebeye. In Majuro, the interventions achieved an estimated 55% reduction in the local TB incidence and mortality in 2020, compared to the counterfactual no intervention scenario, with effects sustained until at least 2050 with no further intervention. In Ebeye, the estimated incidence reduction induced by the screening activities reached 43% in 2050, compared to the counterfactual no intervention scenario. If current programmatic conditions were continued with no future ACF interventions and assuming enhanced TB awareness following the interventions, we predicted that RMI could achieve the 2025 End TB incidence milestone (Figure 4). However, the country would fall short of reaching any TB mortality targets and the 2035 TB incidence target.

**Figure 3.**
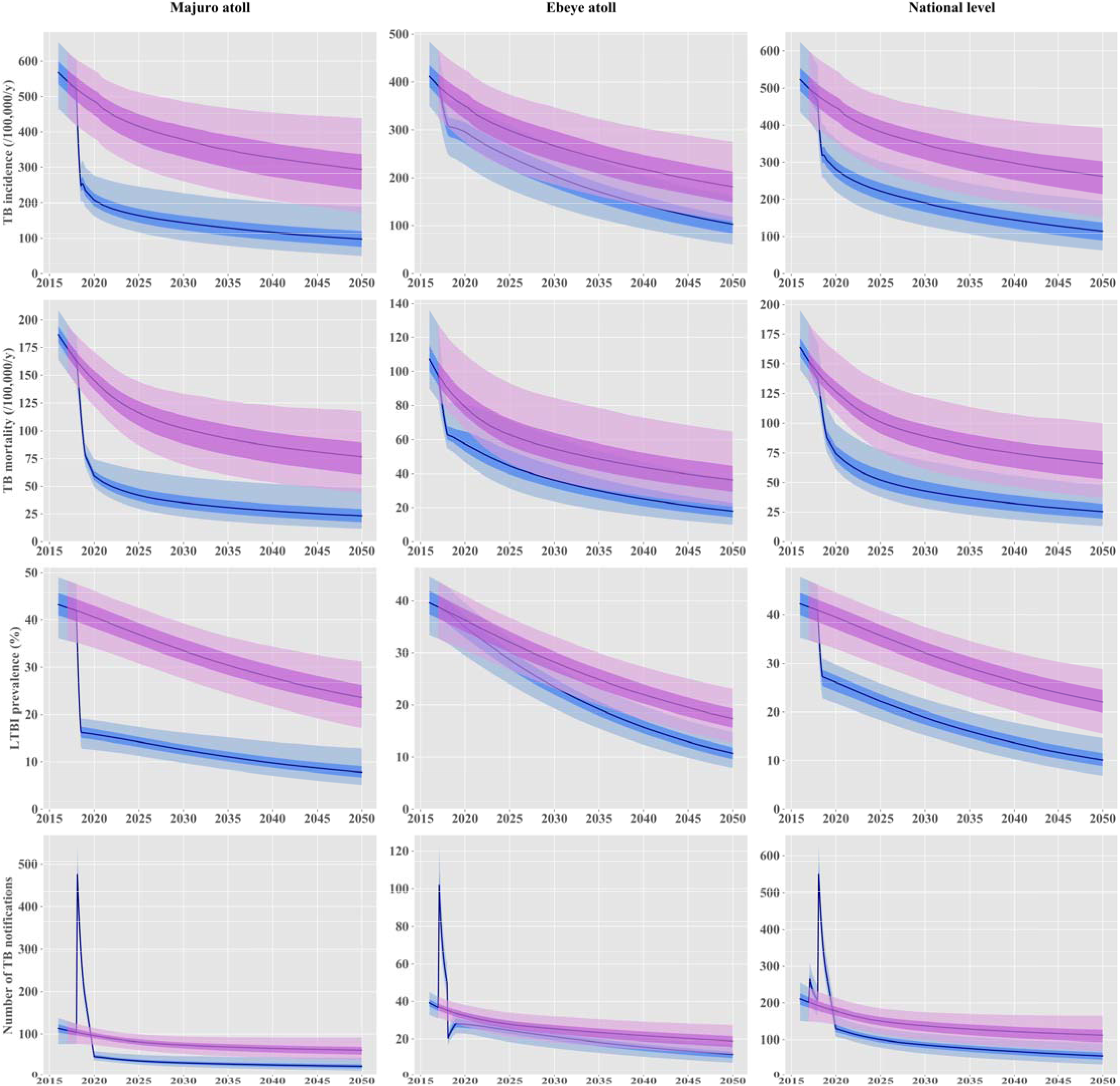
Projected effect of the active screening interventions implemented in 2017 and 2018. The solid lines represent the median estimates. The shaded areas show the interquartile ranges (dark shade) and 95% credible intervals (light shade) projected in the absence of any intervention (pink) and under a scenario including the interventions implemented in 2017-2018 in Majuro and Ebeye (blue).

**Figure 4.**
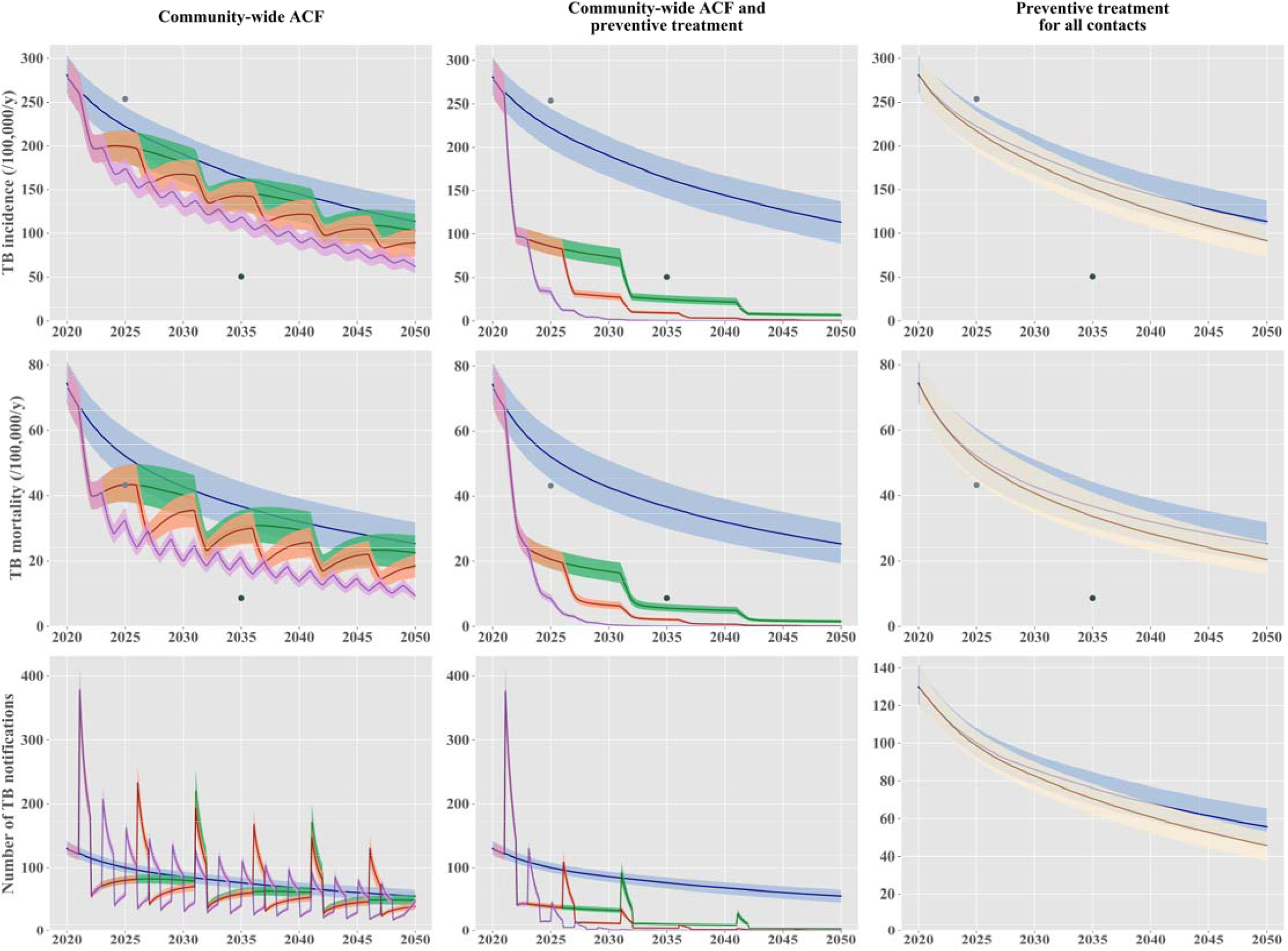
Projected effect of periodic community-wide interventions and contact-tracing-based preventive treatment. The solid lines represent the median estimates, and the shaded areas show the interquartile credible ranges. The “status-quo” scenario is represented in blue in all panels. The left column of panels presents scenarios including nationwide active case finding (ACF) repeated every two years (purple) or every five years (orange) or every ten years (green). The central column of panels presents nationwide ACF scenarios combined with mass latent infection screening and treatment, repeated every two years (purple) or five years (red). The right column of panels shows a hypothetical scenario where all contacts of TB patients would be screened for latent infection and treated if they had a positive test (beige). The light and dark grey dots show the 2025 milestones and the 2035 targets, respectively, according to the End TB Strategy.

The results of our analysis considering different assumptions for the future trend of diabetes prevalence are shown in Supplementary Figure E2. This analysis included the interventions previously conducted in Majuro and Ebeye and assumed that TB control would remain similar to the current programmatic situation until 2050. If diabetes prevalence increased by 20% by 2050, the predicted TB incidence in 2050 would be 140 (95% CrI 73-245) per-100,000-persons-per-year. In contrast, we estimated that a 20% decline in diabetes prevalence would be associated with a further decrease in TB incidence to 92 (95% CrI 51-151) per-100,000-persons-per-year in 2050.

The projections considering repeated interventions at the country level every two, five or ten years are presented in Figure 4. If LTBI screening was not included in the ACF programs, we predicted that the 2035 End TB Strategy targets would not be reached, regardless of the intervention frequency considered. Combining ACF with LTBI screening and treatment would have considerably greater impacts on future TB burden. We estimated that all the End TB Strategy targets would be reached, whether this intervention was repeated every two, five or ten years. Similar results to those presented in Figure 4 are shown on a log-scale in Supplementary Figure E3, displaying the pre-elimination threshold defined as a TB incidence rate of 1 case per-100,000-persons-per-year (15). This showed that RMI could achieve pre-elimination around 2035 with community-wide screening of latent and active TB repeated every two years at the country level, and around 2045 with the same intervention implemented every five years. The ten-year cycle was not predicted to reach pre-elimination by 2050.

Our analysis considering the use of preventive treatment provided through intensive contact tracing showed that such an intervention would yield a comparable overall effect compared to using community-wide ACF at the country level every five years. This intervention would not achieve any of the targets set by the End TB strategy for 2035.

## Discussion

In 2017 and 2018, RMI conducted unprecedented TB and LTBI screening activities. These ambitious community-led interventions were supported by exceptional efforts from local and external stakeholders and supported by volunteers. Success has already been demonstrated through the large number of detected individuals with latent or active TB who completed treatment. Our modelling projections now suggest that these efforts will have considerable effects on the local TB epidemics’ long-term trajectory. We also estimate that periodic use of interventions such as those implemented in RMI could achieve pre-elimination goals over the coming decades.

ACF is a high-intensity, high-resource effort. Going beyond TB case-finding by adding treatment of LTBI increases the cost and the time to screen a population, but our model demonstrates a considerably higher impact when both active and latent TB are addressed together. This suggests that considerable resources are likely to be required to yield the dramatic impacts on TB burden we projected. However, we also show that impacts could be sustained in the long term and that RMI could achieve pre-elimination through such interventions. This means that substantial returns on investment are anticipated as many TB cases and deaths will be averted over several decades. In addition to the human and societal benefits, these direct health effects also translate into economic savings because of the known catastrophic impact that TB has on individuals, families and countries’ finances (16). Our results, therefore, highlight the importance of adopting a long-term vision when planning TB control and when funding the TB response.

Our analysis has important implications for future TB control in RMI. First, we suggest that while it is critical to address latent TB infection, classic approaches relying on tracing the contacts of diagnosed TB cases would be unable to achieve dramatic reductions in burden. An explanation for this is that such strategies would not capture infections that result from old transmission events, whereas these remote infections may still contribute to the future burden of active TB through late reactivation. This suggests that population-wide screening of LTBI may be a necessary component of any program aiming to achieve TB pre-elimination in RMI. The critical importance of reducing the latent infection pool’s size had already been demonstrated by previous modelling works, which suggested that even if transmission ceased from 2015 onwards, this would be insufficient to achieve the End TB Strategy targets globally (17). In the present analysis, we demonstrate that these goals could be achieved by using large-scale screening interventions that are realistic, practical and utilise only existing technologies, given that such interventions have already been implemented in the field. Our study also reinforces the importance of addressing risk factors and comorbidities in addition to TB itself. In particular, we found diabetes to be a key determinant of the TB epidemic’s long-term trends in the RMI context, consistent with findings from other Pacific Island settings (12). This calls for multifactorial approaches that combine TB-specific measures with interventions addressing TB risk factors, whether these tools are medical or social. Finally, our model suggests a clear pathway to reaching TB pre-elimination in RMI. Indeed, we estimated that TB incidence could be brought below one new case-per-100,000-persons-per-year by 2035 if the ambitious intervention conducted in Majuro in 2018 could be repeated every two years from 2021 at the national level.

Our study’s strengths include the fact that our calibration approach was able to capture key disease indicators accurately while incorporating uncertainty around the most fundamental parameters. Furthermore, the model was directly informed by the most relevant data possible since these were directly measured in the field, including through the interventions themselves. This includes data on the prevalence of active and latent TB, which are critical to accurately replicating the local TB burden, along with TB notification data that ensured that the historical trends in case detection were captured appropriately. Finally, we conducted simulations using state-of-the-art computing techniques that are publicly available and have been extensively tested and documented, such that our model could be easily reused to assist TB control in RMI and other settings.

Limitations of our study include the significant uncertainties that remain in TB epidemiology. In particular, model projections could be refined with improved knowledge about the effect of preventive therapy on the risk of future reinfection, although our model considers a broad range of assumptions regarding this parameter. Similarly, even if recent studies have produced estimates for the rates of progression from latent to active TB (13,18), the rate of late reactivation remains poorly characterised due to the limited data available (19). Refining estimates of this parameter would undoubtedly increase the accuracy of predictions related to interventions involving preventive treatment. Second, while our model demonstrates the effects of two different ACF strategies in a high-incidence area, it may not capture all impacts of coordinated TB screening efforts. More nuanced impacts to a local or national TB program could include improvements to TB diagnosis, scaling up efforts to identify close contacts, and increasing resources for LTBI identification and management among contacts and other high-risk groups. These programmatic changes are more difficult to quantify in a way that could be incorporated into a model and could also lead to sustainable TB incidence reductions. Third, the model allows interactions between individuals of different islands, although empiric data were not available to quantify this social mixing process. Therefore, the extent of the diffusion of the interventions’ effects between the different locations relies on our assumption that 95% of interpersonal contacts occur between individuals of the same geographical stratum. Finally, we stress that the projections presented in this study are intended to be primarily relevant to the RMI context and, given the variability of TB epidemics in different settings, drawing quantitative conclusions for other settings would require dedicated analyses (20).

Repeating screening of LTBI within the same population may present some practical issues that are not considered in this modelling study. It can be difficult to interpret positive tests in previously treated individuals since LTBI tests frequently remain positive after treatment.Thus, some individuals may be treated unnecessarily if all positive tests were interpreted as evidence of current infection. Conversely, reinfection occurs, and some infections may be missed if positive tests were systematically interpreted as false positives in previously treated individuals, in which case testing would become redundant for this population. These issues suggest that repeating active screening programs every five years or ten years may be more realistic than two-year cycles. They also highlight the importance of developing highly accurate infection screening tools that could distinguish between previous and current infections or identify biomarkers that could predict short-term progression risk.

In conclusion, our analysis suggests that RMI may not reach all the End TB Strategy targets under the current programmatic situation. However, it would be possible to achieve pre-elimination in the next ten-to-twenty years through periodic repetition of the same interventions as those already implemented in the country, using a similar screening rate at the national level. If this is to occur, it is a rare example of the capability of existing tools to achieve END TB targets.

## Data Availability

The data and code used to implement the study are available on a Github repository.

https://github.com/monash-emu/AuTuMN/tree/master/apps/tuberculosis/regions/marshall_islands

## Acknowledgments

We gratefully thank Dr Richard Brostrom for his detailed description of the interventions, providing access to data, and his advice on capturing the local TB epidemiology accurately. We also thank the nurses, doctors and all persons involved in implementing the screening activities and who contributed to collecting the data used to inform this modelling study. In particular, we thank the late Dr Kennar Briand, Herokko Neamon, Risa Bukbuk and Mareta Hauma for their contribution to this work.

