## Supplement for "Long-term effects of mass screening for latent and active tuberculosis in the Marshall Islands"

*Online Data Supplement*

### Model design

#### Base model

We use a deterministic compartmental model including six types of compartments that represent different states of infection and disease. The model is using the same conceptual approach and similar assumptions to previously published models (E1–E4). Here we describe the model structure before applying any stratification. A susceptible compartment (*S*) is used to represent individuals who have never been infected with *Mycobacterium tuberculosis* (M.tb). Latent TB infection (LTBI) is modelled using two successive compartments: early latent (*E*) and late latent (*L*) to capture the declining risk of disease progression over time from infection (E5). The active disease compartment (*I*) represents individuals who have progressed to the active stage of TB disease. Diseased individuals who recover through self-cure progress directly to the recovered compartment (*R*). All diseased individuals who are detected are assumed to be started on treatment (compartment *T*). Treatment may result in cure (progression to *R*), relapse (return to *I*) or death.

Non-TB-related mortality is modelled by applying death rates to all model compartments. In addition, disease-specific mortality is implemented by applying increased mortality rates to the active disease compartments (*I* and *T*).

Reinfection occurs in the model in two different ways. First, latently infected individuals may be reinfected, and this process is modelled using a flow from the late latent (*L*) to the early latent compartment (*E*). Second, individuals who have recovered from TB disease may be reinfected and return to the early latent compartment. The structure of our model allows for differential risk of infection for the currently and previously infected individuals, compared to the infection-naive individuals.

Figure 1 (main text) represents the model structure.

#### Stratification by organ status

The model explicitly includes three types of TB clinical manifestations, based on the organ affected by the disease and the smear status. The three “organ categories” are smear-positive TB, smear-negative pulmonary TB and extrapulmonary TB. The term “smear-negative TB” will be used to refer to “smear-negative pulmonary TB” hereafter.

The stratification by organ status applies to the active disease compartments (*I* and *T*), and we assume differential disease fatality rates, infectiousness levels and detection rates by organ status.

#### Stratification by age

The model is stratified using five categories: 0-4, 5-14, 15-34, 35-49 and 50+ years old. We assume heterogeneous mixing by age using age-specific contact matrices published by Prem and colleagues (E6). We used estimates reported for Kiribati as a proxy for the Marshall Islands since no local estimates were available. The matrices were calculated using 16 age groups (“0-4”, “5-9”, ..., “70-74”, “75+”) in the original study. In the model, the original 16-by-16 matrix is converted to a 5-by-5 matrix to match our model stratification. This conversion is done by aggregating the contact rates across the relevant age groups and applying weighted averages based on the population age distribution.

#### Stratification by location

All model compartments are further stratified by location to explicitly represent the respective populations of the Majuro Atoll, the Ebeye Island and other islands. Population proportions by location were informed by the 2011 National Census. We assumed that individuals make 95% of their contacts with persons living in the same geographic stratum. The remaining 5% are evenly distributed between the two other locations. The three locations shared the same values for most model parameters, but we allowed for differential passive screening rates by location, and the interventions were only implemented in the Ebeye Atoll (ACF) and in the Majuro Atoll (ACF and LTBI screening).

Table E1 summarises the different stratifications implemented in the model and highlights the key changes applied to the stratified parameters.

| **Stratification** | **Strata** | **Significance to model** |
| --- | --- | --- |
| Age | 0-4 years old  5-14 years old  15-34 years old  35-49 years old  50 years and over | - Risk of progression from latent to active TB varies with age. - Background mortality rates vary with age. - Age-specific infectiousness. - Diabetes prevalence varies with age. - Heterogenous mixing by age - BCG vaccine effect and coverage varies with age. |
| Location | Majuro  Ebeye  Other | - Heterogenous mixing between the three geographical groups will simulate the impact of inter-island travel on the population effect of the interventions in Ebeye and Majuro. 95% of social contacts occur with individuals from the same region. 2.5% with individuals from each of the other two regions. - Case detection rates may vary between the regions - The rates of LTBI screening and active case finding vary by location |
| Organ status | Pulmonary smear-positive  Pulmonary smear-negative  Extrapulmonary | - Case detection rates vary according to organ status. - Smear-positive TB has a higher level of infectiousness than smear-negative TB, and extrapulmonary TB is considered non-infectious. |

**Table E1. Summary of model stratifications**

#### Births and deaths

Births are modelled using time-variant crude birth rates that are multiplied by the modelled population size to determine the number of newborn individuals entering the model at each time. A time-variant and age-specific rate or non-TB-related mortality applies to all model compartments to simulate deaths from other causes than TB. We use estimates from the UN population division to inform the birth and mortality rates. No data specific to the Marshall Islands were available, so we used the crude birth rates and mortality rates by age of the Federated States of Micronesia.

We also apply additional death rates to the compartments *I* and *T* to reflect mortality induced by TB disease.

#### *M.tb* transmission

We use different levels of susceptibility to infection for individuals who are currently latently infected with M.tb or have recovered from active TB, as compared to infection-naive individuals. The effect of BCG vaccination is captured by reducing the susceptibility to infection of individuals under the age of 30 years old. We assume a 70% reduction in the susceptibility of BCG-vaccinated children under the age of 15 years old (E7). A linear function is used to reflect the progressive loss of BCG immunity between the age of 15 and 30 years old. Figure E1 presents the profile of BCG immunity wane. The continuous wane profile is then automatically converted to a step function such that each model’s age band is associated with the average value of the wane function over the relevant interval. The susceptibility adjustment induced by BCG is also based on the country’s time-variant BCG coverage as reported by the WHO. The rest of this section describes the different infectiousness adjustments implemented in the model.

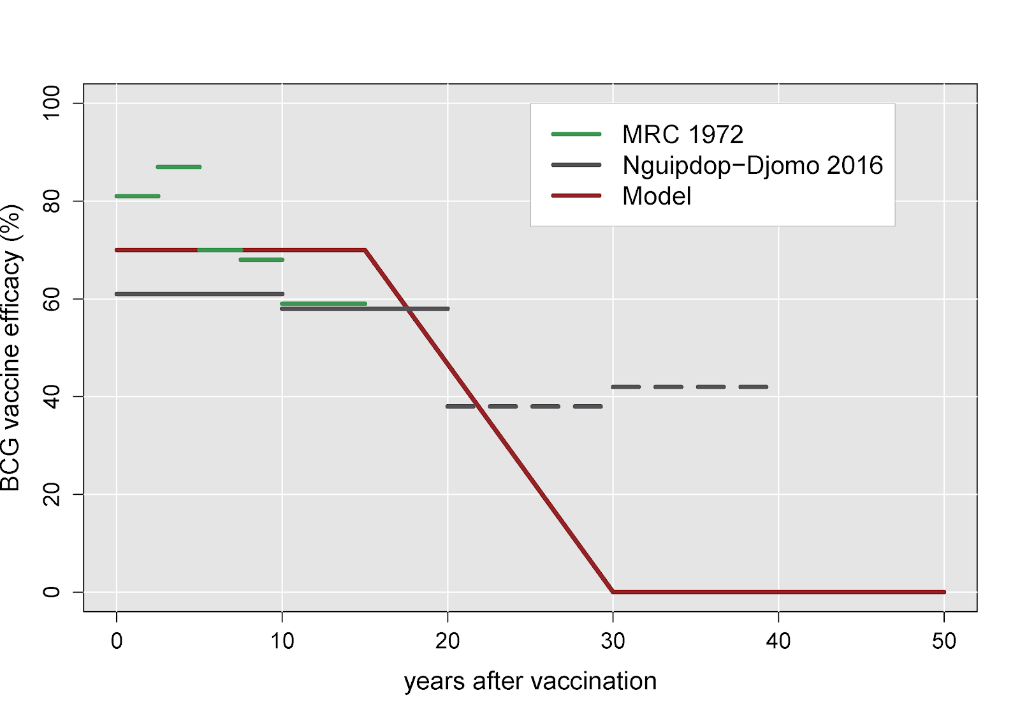

**Figure E1. Assumed wane profile of BCG efficacy.**

Green and grey lines represent estimates obtained from literature (E7,E8), while the red line shows the modelled vaccine effect. Dashed lines show estimates associated with non-significant efficacy.

We assume that smear-negative TB is 25% as infectious as smear-positive TB, while extrapulmonary TB is modelled as a non-infectious disease state (E9,E10).

Infectiousness is assumed higher for older individuals, and we use the sigmoidal function $age\to\frac{1}{1+e^{-(age-15)}}$ to model a progressive increase with age (E11). The continuous age profile of infectiousness is then automatically converted to a step function, similarly to what was previously described for the immunity wane profile of BCG vaccination.

Finally, individuals who are on treatment are assumed to be partially infectious, as infectiousness declines rapidly after treatment initiation. Infectiousness is multiplied by 0.08 for the treatment compartment compared to the untreated disease compartment to reflect the fact that individuals may remain infectious for two weeks out of the 26 weeks of a standard regimen.

### Progression parameters

#### Progression from latent to active TB

We use the estimates reported in Ragonnet *et al.* to inform the modelled dynamics of activation from latent to active TB (Table 2, main text) (E5). These parameters vary by age, and a multiplier is used to incorporate uncertainty around the progression rates (E5).

#### Effect of diabetes

The model is not stratified by diabetes status. Instead, we model the effect of diabetes type 2 by increasing the rates of progression from latent to active TB using age-specific multipliers. For each age group, the value of the diabetes-effect multiplier depends on the age-specific proportion of diabetic individuals and the relative rate of TB reactivation for diabetic individuals compared to non-diabetic individuals (see Table 2, main text). The updated TB progression rates can be written $m_{i}=d_{i}\times rr_{diabetes}+1-d_{i}$, where $d_{i}$ is the proportion of diabetic individuals in the age-group $i$ and $rr_{diabetes}$ is the relative rate of TB reactivation for diabetic individuals compared to non-diabetic individuals.

Diabetes prevalence was assumed to increase progressively to reach the values presented in Table 2 in 2020 (main text) and was then assumed to be constant after that time in the base case analysis. However, we considered both a linear increase of 20% and a linear decrease of 20% by 2050 in sensitivity analyses.

#### Natural history flows

We use the estimates reported in Ragonnet *et al.* to model the rate of TB mortality in the absence of treatment and the rate of self-recovery (E12). We use different rates of untreated TB mortality and self-recovery for smear-positive TB compared to smear-negative TB. The TB mortality and self-recovery rates associated with extrapulmonary TB are assumed to be the same as those of smear-negative TB.

#### Passive detection of active TB

The detection rate is defined as the rate of progression from the active disease to the treatment compartment, as all detected individuals are assumed to be started on treatment at diagnosis in our model. This rate is calculated by multiplying the screening rate with the diagnostic test sensitivity. The screening rate can be interpreted as the reciprocal of the average time that diseased individuals take to seek care. The diagnostic sensitivity varies according to the organ status to reflect the relative differences in the difficulty to diagnose smear-negative TB and extrapulmonary TB, as compared to smear-positive TB.

We use a time-variant function to model the screening rate in order to capture detection improvements over time. This process is modelled assuming a continuous increase in the screening rate through the following function:

$t\to0.5(\tanh\left( m\left( t-c \right)+1 \right)\times final$, where $c$ is the time at which the curve inflects, $m$ is the value of the gradient at the inflection point (shape parameter), and $final$ is the upper asymptote value. The parameters $c$, $final$ and $m$ are varied during calibration, and Figure E2 presents the time-variant profile of screening rate obtained by sampling these parameters from their posterior joint distribution.

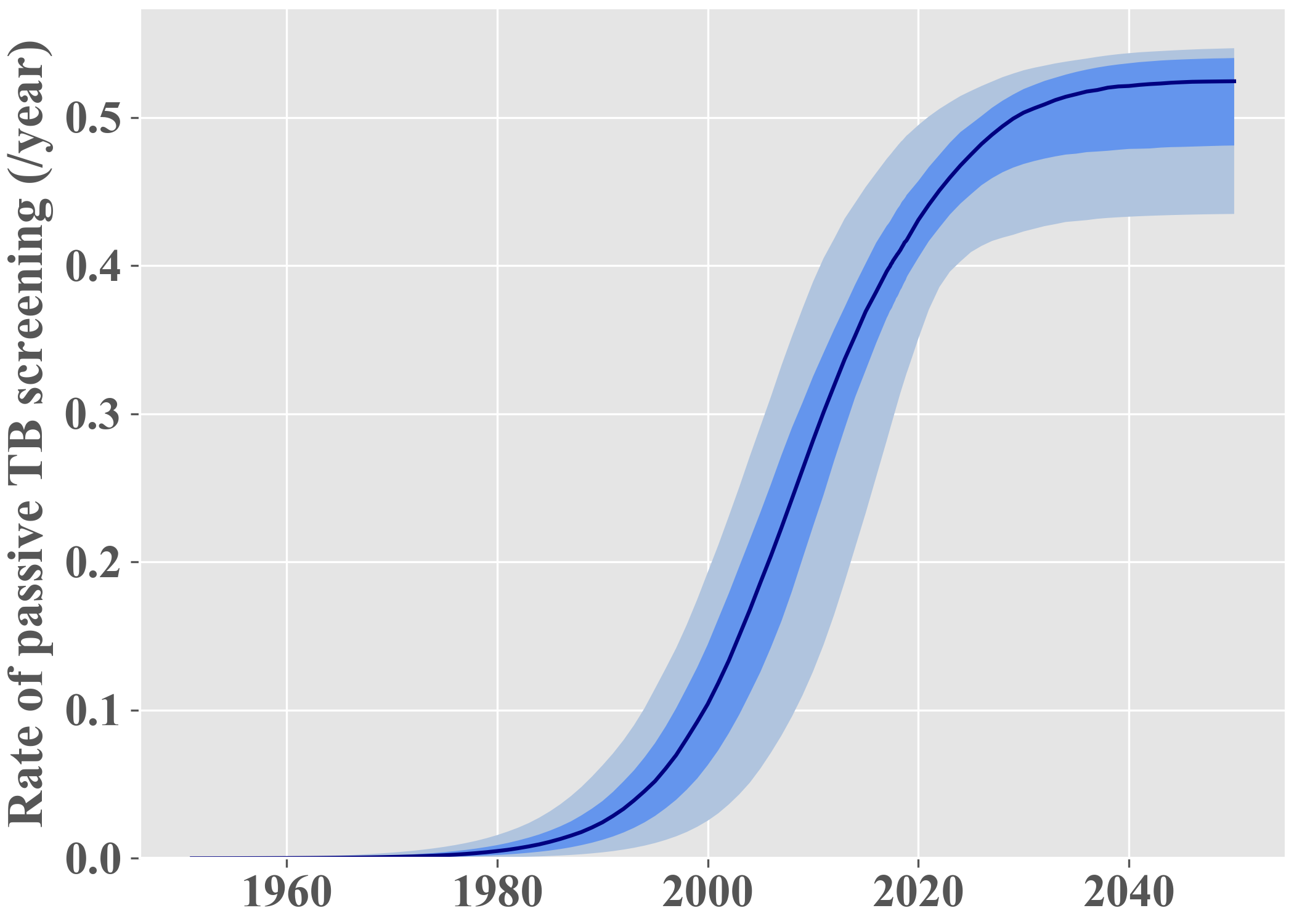

**Figure E2. Posterior estimate of the passive screening rate profile**

Green and grey lines represent estimates obtained from literature (E7,E8), while the red line shows the modelled vaccine effect. Dashed lines show estimates associated with non-significant

#### Treatment outcomes

Treated individuals can experience three different treatment outcomes: treatment success, relapse or death. The rate of treatment-induced recovery ($\varphi$) is set to the reciprocal of the duration of a completed treatment course. We then use the observed treatment success proportion (often referred to as “treatment success rate”) as model input. In our model, it is calculated from $TSR=\frac{\varphi}{\varphi+\rho+\mu_{T}+\mu}$, where $\rho$ is the relapse rate, $\mu_{T}$ is the excess mortality rate of individuals on TB treatment, and $\mu$ is the non-TB-related mortality rate. Finally, we calculate the respective values of $\rho$ and $\mu_{T}$ using the observed proportion of deaths among all negative treatment outcomes, denoted $\pi$. We have $\pi=\frac{\mu_{T}+\mu}{\rho+\mu_{T}+\mu}$ that we inject into the $TSR$ equation. After calculations, we obtain:

$\mu_{T}=\pi\varphi\frac{1-TSR}{TSR}-\mu$ and $\rho=(\mu_{T}+\mu)(\frac{1}{\pi}-1)$.

Note that we need to verify $\varphi\geq\frac{\mu}{\pi}\times\frac{TSR}{1-TSR}$ to ensure that $\mu_{T}\geq0$. If this condition is not verified, which may be the case if both the treatment success rate and the rate of non-TB-related mortality are high, we force $\varphi= \frac{\mu}{\pi}\times\frac{TSR}{1-TSR}$.

Since the non-TB-related mortality rate varies by age, the values of the treatment-induced recovery ($\varphi$), relapse rate ($\rho$) and mortality rate of individuals on TB treatment ($\mu_{T}$) also vary by age.

### Modelled interventions

#### Population-based screening and treatment of LTBI

Mass LTBI screening and treatment is implemented as part of the intervention conducted in Majuro in 2018. This is modelled by making latently infected individuals (from *E* and *L*) transition to the recovered compartment (*R*). The rate associated with these flows is obtained by multiplying the LTBI screening rate with the sensitivity of the LTBI test employed and the individual-level efficacy of preventive treatment. The LTBI screening rate is implemented as a time-variant parameter that is stratified by location.

#### Active case finding

Active case finding (ACF) is implemented to simulate the interventions linked to the detection of individuals with active TB implemented in Ebeye in 2017 and in Majuro in 2018. This is modelled by implementing an additional transition flow from compartment *I* to compartment *T*. The rate associated with this flow is obtained by multiplying the location-specific ACF screening rate with the sensitivity of the detection algorithm used for the ACF intervention. The ACF screening rate is implemented as a time-variant parameter.

#### Calculation of the screening rates

To simulate the interventions, we apply a positive rate of ACF and/or LTBI screening over the intervention periods. The screening rates are determined such that the modelled total proportion of the population screened corresponds to the true population proportion screened. The screening rate is set equal to $-log(1-coverage)$ for the year during which the intervention is implemented, where $coverage$ is the total proportion of the population screened by the intervention. In the Ebeye Atoll, it was estimated that 85% of adult individuals (aged 15 years old and over) were screened for active TB. In the Majuro Atoll, 22,623 individuals out of a population of 27,797 population were screened for both LTBI and active TB ($coverage=81\%$). The screening rate obtained from the Majuro intervention was used to model the intervention scenarios involving country-wide screening repeated periodically.

### Model calibration

#### Overall approach

The model is calibrated using an adaptive Metropolis (AM) algorithm. We use the multidimensional Gaussian distribution with variable covariance matrix presented by Haario and colleagues to sample parameters from their posterior distributions (E13). We ran seven independent AM chains initialised using Latin Hypercube Sampling across the uncertainty parameters being calibrated. The number of iterations is limited by a maximum computational time of 5 hours. We discarded the first 500 iterations of each chain as burn-in and combined the samples of the seven chains to project epidemic trajectories over time.

The likelihood function was derived from comparing model outputs to target data at each time point nominated for calibration. We used normal distributions centred on the model predictions, and the standard deviations of these distributions were considered as calibration parameters.

#### Parameters’ prior distributions

We use uniform prior distributions characterised by the intervals presented in Table 2 (main text) and Table E2 below.

| **Parameter** | **Range** |
| --- | --- |
| Initial population size | 200 - 800 |
| Transmission scaling factor | 0.2 - 1.0 |
| Progression multiplier | 0.5 - 2.0 |
| Screening profile (inflection time), year | 2000.0 - 2020.0 |
| Screening profile (shape) | 0.07 - 0.1 |
| Screening profile (final rate), per year | 0.4 - 0.55 |
| Relative rate of passive TB screening in Ebeye (ref. Majuro) | 1.3 - 2.0 |
| Relative rate of passive TB screening in other islands (ref. Majuro) | 0.5 - 1.5 |
| Relative rate of TB progression for diabetic individuals | 2.0 - 5.0 |
| Relative risk of infection for individuals with latent infection (ref. Infection-naive) | 0.2 - 0.5 |
| Relative risk of infection for individuals with history of infection (ref. Infection-naive) | 0.2 - 1.0 |
| Efficacy of preventive treatment | 0.75 - 0.85 |
| Relative screening rate following ACF interventions (ref. Before intervention) | 1.0 - 1.5 |
| TB mortality (smear-positive), per year | 0.335 - 0.449 |
| TB mortality (smear-negative), per year | 0.017 - 0.035 |
| Self-cure rate (smear-positive), per year | 0.177 - 0.288 |
| Self-cure rate (smear-negative), per year | 0.073 - 0.209 |

**Table E2. Prior distribution ranges**

#### Calibration targets

The following table summarises the calibration targets used to calculate the likelihood of the AM algorithm.

| **Variable** | **Targeted value** | **Source** |
| --- | --- | --- |
| TB prevalence in Majuro in 2018 | 1366 per 100,000 persons | Measured during intervention |
| TB prevalence in Ebeye in 2017 | 755 per 100,000 persons | Measured during intervention |
| LTBI prevalence in Majuro in 2018 | 38% | Measured during intervention, adjusted for test sensitivity. |
| TB notifications in Majuro in   - 2012 - 2013 - 2014 - 2015 - 2016 - 2017 - 2018 - 2019 - 2020 | - 91 - 116 - 115 - 90 - 119 - 116 - 376 - 135 - 96 | TB program |
| TB notifications in Ebeye in   - 2012 - 2013 - 2014 - 2015 - 2016 - 2017 - 2018 - 2019 - 2020 | - 44 - 27 - 30 - 29 - 53 - 80 - 30 - 45 - 39 | TB program |
| Total population size in 2011 | 53158 | 2011 National Census |

**Table E3. Calibration targets**

#### Posterior estimates of calibrated parameters

Table E4 presents the posterior estimates obtained for the fitted parameters. The percentiles were obtained after combining the samples from the seven AM chains.

| **Parameter** | **2.5th percentile** | **Median** | **97.5th percentile** |
| --- | --- | --- | --- |
| Initial population size | 335 | 522 | 737 |
| Transmission scaling factor | 0.444 | 0.599 | 0.792 |
| Progression multiplier | 1.45 | 1.79 | 1.99 |
| Screening profile (inflection time), year | 2000 | 2010 | 2020 |
| Screening profile (shape) | 0.0709 | 0.0823 | 0.0967 |
| Screening profile (final rate), per year | 0.435 | 0.525 | 0.547 |
| Relative rate of passive TB screening in Ebeye (ref. Majuro) | 1.47 | 1.83 | 1.99 |
| Relative rate of passive TB screening in other islands (ref. Majuro) | 0.704 | 1.22 | 1.46 |
| Relative rate of TB progression for diabetic individuals | 4.6 | 4.95 | 5 |
| Relative risk of infection for individuals with latent infection (ref. infection-naive) | 0.203 | 0.345 | 0.479 |
| Relative risk of infection for individuals with history of infection (ref. infection-naive) | 0.246 | 0.58 | 0.905 |
| Efficacy of preventive treatment | 0.756 | 0.801 | 0.849 |
| Relative screening rate following ACF interventions (ref. before intervention) | 1.05 | 1.3 | 1.48 |
| TB mortality (smear-positive), per year | 0.335 | 0.337 | 0.38 |
| TB mortality (smear-negative), per year | 0.017 | 0.0196 | 0.0304 |
| Self-cure rate (smear-positive), per year | 0.178 | 0.184 | 0.256 |
| Self-cure rate (smear-negative), per year | 0.073 | 0.073 | 0.0896 |

**Table E4. Parameter posterior estimates**

### Sensitivity analysis varying the future prevalence of diabetes

The results of our analysis considering different assumptions for the future trend of diabetes prevalence are shown in Figure E3. This analysis included the interventions previously conducted in Majuro and Ebeye and assumed that TB control would be similar to the current programmatic situation until 2050. We considered both a linear increase of 20% (red line and shade) and a linear decrease of 20% (green line and shade) in diabetes prevalence between 2020 and 2050 in this sensitivity analysis.

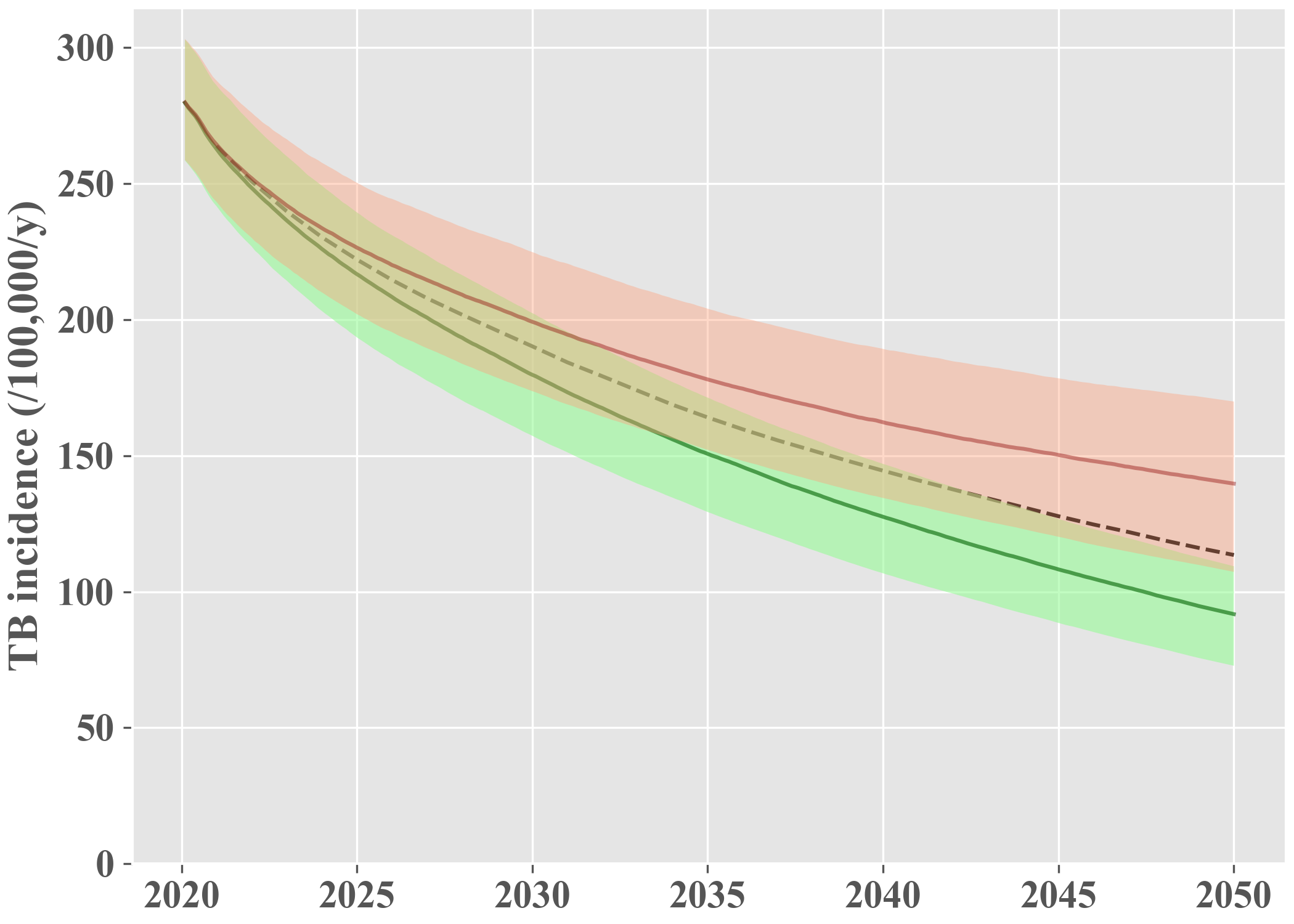

**Figure E3. Projected epidemic under status-quo scenario considering different future trends for diabetes prevalence.**

The dashed line represents the base case median estimate associated with a constant future diabetes prevalence, maintained at the same level as in 2020. The green (resp. red) line shows the median projection associated with a 20% decrease (resp. increase) in diabetes prevalence by 2050. The shades represent the interquartile ranges.

### Projected impact of periodic interventions

The projected epidemics under the different intervention scenarios are presented in the main text (Figure 4). Here below, the same outputs are represented on a log scale in order to visualise whether the interventions considered could achieve the pre-elimination target by 2050.

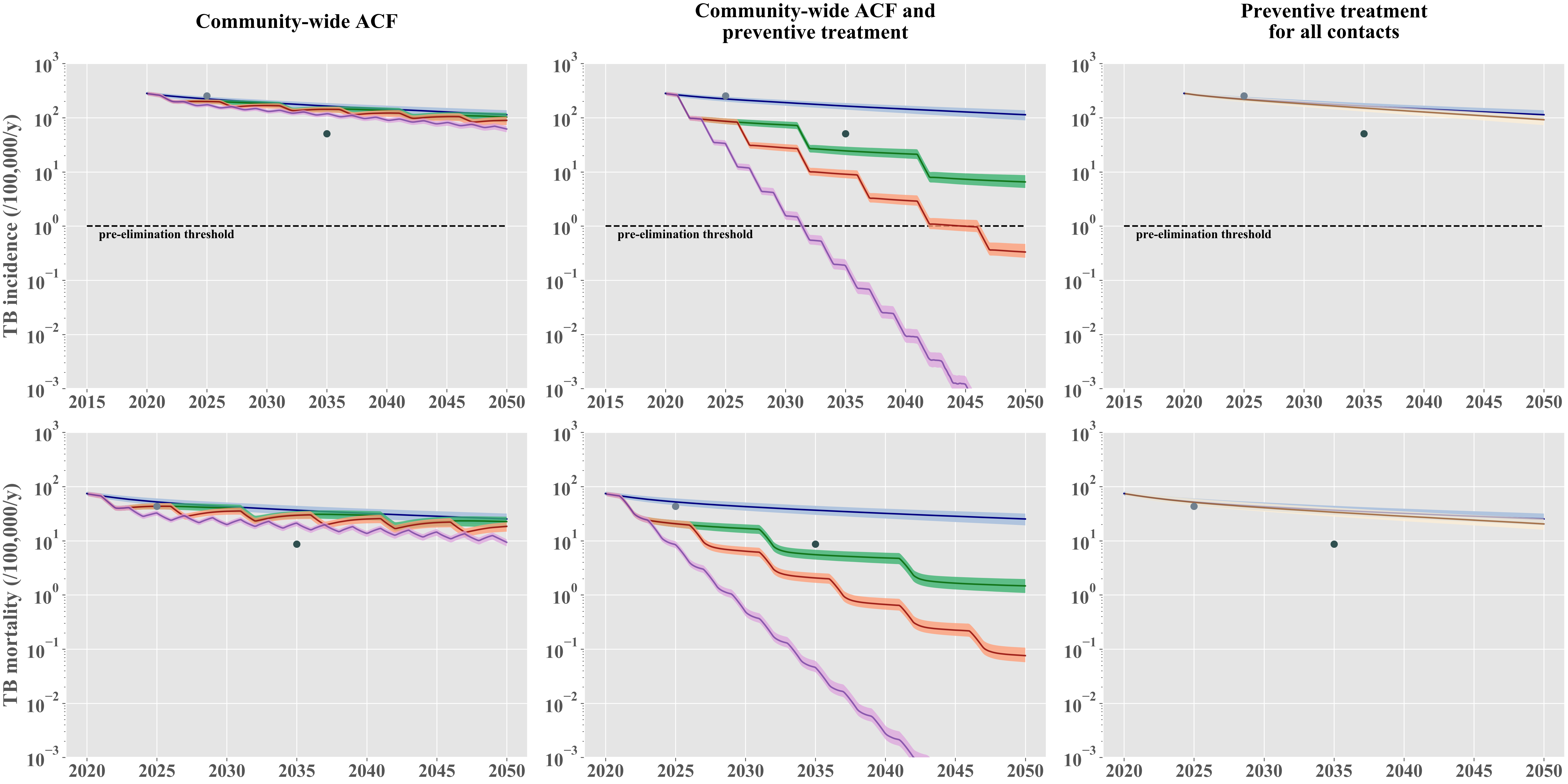

**Figure E4. Projected effect of periodic community-wide interventions and contact-tracing-based preventive treatment (log-scale).**

The solid lines represent the median estimates, and the shaded areas show the interquartile ranges. The “status-quo” scenario is represented in blue in all panels. The left column presents scenarios including nationwide active case finding (ACF) repeated every two years (purple), every five years (orange), or every ten years (green). The central column presents scenarios including nationwide ACF combined with mass latent infection screening and treatment, repeated every two years (purple) or five years (red). The right column shows a hypothetical scenario where all contacts of TB patients would be screened for latent infection and treated if they had a positive test. The light and dark grey dots show the 2025 milestones and the 2035 targets, respectively, according to the End TB Strategy. The horizontal dashed lines represent the pre-elimination threshold defined by the WHO as a TB incidence of 1 per-100,000-persons-per-year.

E5.	Ragonnet R, Trauer JM, Scott N, Meehan MT, Denholm JT, McBryde ES. Optimally capturing latency dynamics in models of tuberculosis transmission. Epidemics. 2017;

E6.	Prem K, Cook AR, Jit M. Projecting social contact matrices in 152 countries using contact surveys and demographic data. PLoS Comput Biol [Internet]. 2017;13(9):e1005697. Available from: http://www.ncbi.nlm.nih.gov/pubmed/28898249

E7.	Nguipdop-Djomo P, Heldal E, Rodrigues LC, Abubakar I, Mangtani P. Duration of BCG protection against tuberculosis and change in effectiveness with time since vaccination in Norway: a retrospective population-based cohort study. Lancet Infect Dis [Internet]. 2016;16(2):219–26. Available from: http://www.ncbi.nlm.nih.gov/pubmed/26603173

E8.	Medical Research Council. BCG and vole bacillus vaccines in the prevention of tuberculosis in adolescence and early adult life. Bull World Heal Organ [Internet]. 1972/01/01. 1972;46(3):371–85. Available from: http://www.ncbi.nlm.nih.gov/pubmed/4537855

E9.	Behr MA, Warren SA, Salamon H, Hopewell PC, Ponce de Leon A, Daley CL, et al. Transmission of Mycobacterium tuberculosis from patients smear-negative for acid-fast bacilli. Lancet [Internet]. 1999;353(9151):444–9. Available from: http://www.ncbi.nlm.nih.gov/pubmed/9989714

E10.	Tostmann A, Kik S V, Kalisvaart NA, Sebek MM, Verver S, Boeree MJ, et al. Tuberculosis transmission by patients with smear-negative pulmonary tuberculosis in a large cohort in the Netherlands. Clin Infect Dis [Internet]. 2008;47(9):1135–42. Available from: http://www.ncbi.nlm.nih.gov/pubmed/18823268

E11.	Ragonnet R, Trauer JM, Geard N, Scott N, McBryde ES. Profiling Mycobacterium tuberculosis transmission and the resulting disease burden in the five highest tuberculosis burden countries. BMC Med [Internet]. 2019 Nov 22 [cited 2020 May 16];17(1):208. Available from: https://bmcmedicine.biomedcentral.com/articles/10.1186/s12916-019-1452-0

E12.	Ragonnet R, Flegg JA, Brilleman SL, Tiemersma EW, Melsew YA, McBryde ES, et al. Revisiting the Natural History of Pulmonary Tuberculosis: A Bayesian Estimation of Natural Recovery and Mortality Rates. Clin Infect Dis. 2020;

E13.	Haario H, Saksman E, Tamminen J. An adaptive Metropolis algorithm. Bernoulli. 2001;
